# Comparing the risks of environmental carcinogenic chemicals in Japan using the loss of happy life expectancy indicator

**DOI:** 10.1101/2023.02.27.23286547

**Authors:** Michio Murakami, Kyoko Ono, Yoshitake Takebayashi, Masaharu Tsubokura, Shuhei Nomura

## Abstract

In this study, we aimed to use the loss of happy life expectancy (LHpLE), an indicator that enables risk assessment considering wellbeing, to compare the risks of environmental carcinogenic chemicals in Japan. First, we surveyed Japanese people to determine their emotional happiness by age and sex and evaluated whether cancer incidence reduced emotional happiness. Questionnaires were administered to a general population panel and a panel of patients with cancer in 2022, recruiting a predetermined number of responses of 5000 and 850, respectively. Second, using the survey data, LHpLE was calculated for radon, arsenic, and fine particulate matter (aerodynamic diameter <2.5 μm; PM_2.5_) and compared to psychological distress, considering increased mortality and decreased emotional happiness due to these risks. We discovered no significant decrease in emotional happiness due to cancer incidence and no significant associations between emotional happiness and cancer type, history, or stage. LHpLE was calculated to be 6.4 × 10^−3^ years for radon, 2.6 × 10^−3^ years for arsenic, 1.1 × 10^−2^ years (2012 exposure) and 8.6 × 10^−4^ years (2020 exposure) for PM_2.5_, and 9.7 × 10^−1^ years for psychological distress. The fraction of losses caused by these carcinogenic chemicals to HpLE exceeded 10^−5^, suggesting that risk reduction for these chemicals is important in environmental policies. The LHpLE indicator allows for comparing different types of risks, such as environmental chemicals and psychological distress. This is the first study to compare chemical risks using the LHpLE indicator.

## 1. Introduction

Comparing risks is the foundation for rational decision-making. Risk comparisons are useful for individual decision-making through an understanding of risk magnitude and social decision-making, such as identifying public health priorities and evaluating risk trade-offs. Common indicators must be utilized to compare the different risks. Examples of risk indicators include mortality (Graunt, 1662), cancer risk (Mantel and Bryan, 1961), loss of life expectancy (LLE; average shortened life expectancy) (Cohen and Lee, 1979), disability-adjusted life years (DALYs) (Murray et al., 1994), and quality-adjusted life years (Zeckhauser and Shepard, 1976). Comparing the risks of environmental chemicals (Gamo et al., 2003) and multinational comparisons of risk changes over time (GBD 2019 Risk Factors Collaborators, 2020) are typical examples of such practices. These risk comparisons can provide information for advancing effective policies and evaluating their effectiveness.

Recently, the importance of considering improvements in wellbeing in environmental policies has been highlighted (Bok, 2010), and evaluations based on wellbeing have been conducted (Fisher et al., 2021; Sanderson et al., 2013). Developing a risk-comparison indicator that considers wellbeing has also been proposed (Johnson et al., 2016). Murakami et al. (2018) combined life tables and subjective emotional happiness based on “yes/no” responses to a questionnaire to determine happy life expectancy (HpLE) and proposed a novel risk indicator for the loss of HpLE (LHpLE) as the average shortened HpLE time. In their study, HpLE was defined as the lifespan during which a person experiences subjective emotional wellbeing. LHpLE can be evaluated by considering mortality and changes in emotional happiness due to risk exposures. As an extended concept of LLE, DALYs, and quality-adjusted life years, LHpLE can be used in various fields, including environmental science, medicine, and economics. LHpLE has several advantages: it is based on a ratio scale; it can be treated arithmetically; it does not require the evaluation of weights related to the trade-off between disability and death; and it can evaluate qualities other than disability. We have compared the psychological distress and cancer risk associated with radiation exposure after the Fukushima Daiichi Nuclear Power Station accident (Murakami et al., 2018), the benefits of returning to one’s hometown after the accident, cancer risks associated with radiation exposure (Murakami et al., 2021), comfort benefits from air conditioning use, and increased risk of heat stroke and flooding associated with climate change (Arai et al., 2020). However, the LHpLE indicator has not been applied to environmental chemicals. Furthermore, changes in emotional happiness due to cancer have not been adequately investigated. Examining whether cancer incidence reduces emotional happiness, applying LHpLE to environmental carcinogenic chemicals, and comparing these to other medical risks can provide a basis for environmental policy prioritization and policy evaluation based on risk reduction over time.

Therefore, we aimed to use the LHpLE, an indicator that enables risk assessment considering wellbeing, to compare the risks of environmental carcinogenic chemicals in Japan. First, we conducted a questionnaire survey of Japanese people to determine their emotional happiness by age and sex and evaluate whether cancer reduces emotional happiness. Second, LHpLE was calculated for radon, arsenic, and fine particulate matter (aerodynamic diameter <2.5 μm; PM_2.5_), which are typical environmental carcinogenic chemicals in Japan, and compared their risk with psychological distress risk, which decreases emotional happiness. We chose psychological distress as a public health risk because it differs from these carcinogens. A large increase in using the LHpLE indicator for psychological distress was observed after the Fukushima Daiichi Nuclear Power Station accident (Murakami et al., 2018); however, psychological distress risk during normal times is unknown. This is the first study to compare chemical risks using the LHpLE indicator.

## 2. Methods

### 2.1. Ethics

The study was approved by the Research Ethics Committee of the Graduate School of Medicine, The University of Tokyo (authorization number 2021318NI-(1)). The participants’ consent was obtained for questionnaire surveys.

### 2.2. Questionnaire surveys

Two online surveys were conducted, one each for the general and disease panels with patients with a cancer history in Japan. These panels were managed by Cross Marketing Inc., along with the health history registration information. Cross Marketing is a large research company with approximately 3 million active panel monitors who responded to surveys within the last year (as of January 2022). Respondents were incentivized with points that could be converted into products or services.

Our first survey was conducted between July 12 and 13, 2022, with the general panel of monitors aged 20–69 until a pre-designed target collection number of 5,000 was achieved. The target collection numbers were set by age and sex to align with age and sex distribution strata in Japan, that is, based on a quota sampling method. Participants were first asked about their consent, age, and sex. Subsequently, participants were asked the following questions: positive wellbeing (enjoyment: “Did you experience a feeling of enjoyment yesterday? [yes, no],” emotional happiness: “Did you experience a feeling of happiness yesterday? [yes, no],” and laughter: “Did you laugh yesterday? [yes, no]”) (Kahneman and Deaton, 2010; Murakami et al., 2020; Murakami et al., 2018), subjective feelings of health, psychological distress (Kessler et al., 2003), health history, including hypertension, diabetes, hyperlipidemia, and cancer, job, marital status, presence/absence of children, presence/absence of grandchildren, educational background, presence/absence of a jobless person within the household, and smoking habits. Enjoyment, emotional happiness, and laughter are positive wellbeing items, and participants were asked to validate emotional happiness in this study. Survey items other than wellbeing and cancer history were used as covariates to assess whether there was a cancer-related decline in emotional happiness, in line with a previous study (Murakami et al., 2018).

Our second survey was administered to those aged 20 years and older with a history of cancer in the disease panel between September 1 and 2, 2022, until the pre-designed target collection number of 850 participants was reached. The quota sampling method was not applied to the second survey. Participants were first asked about their consent and health history, including cancer and age. Participants were asked to complete the same questionnaire items as in the general panel. In addition, they were asked about cancer-related survey items (cancer type, history, and stage).

### 2.3. LHpLE estimation

The details of the calculation of HpLE and LHpLE have been described in a previous study (Murakami et al., 2018). Briefly, life expectancy *e*_*x,s*_ for a given age *x* and sex *s* (males or females) was calculated using the 2010 Japanese life table (Ministry of Health Labour and Welfare, 2012). *HpLE*_*x,s*_ for a given age *x* and sex *s* was calculated using age- and sex-specific values of emotional happiness, as in eqs. 1 and 2 below.

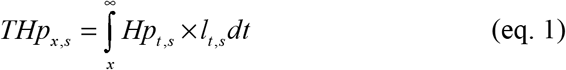

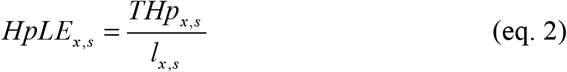

where *Hp*_*x,s*_ is emotional happiness at age *x* and sex *s*, and *l*_*x*_ is the number of people living at the start of an interval between age *x* and sex *s* (of 100,000 born alive).

*Hp*_*x,s*_ is based on the proportion of emotional happiness for each age and sex group (20s, 30s, 40s, 50s, and 60s) for the general panel. The values of 20s and 60s were used for those aged 19 years and younger and for those aged 70 years and older, respectively.

LHpLE is calculated for risk exposures that result in decreased emotional happiness or increased mortality. *LHpLE*_*x,s*_ for a given age *x* and sex *s* was calculated using the difference in *HpLE*_*x,s*_ with and without risk exposure. Unless otherwise noted, HpLE and LHpLE refer to their respective values at age 0. In addition, we calculated the fraction of losses caused by risk exposures to HpLE (*Fr*_*s*_) using eq. 3 below.

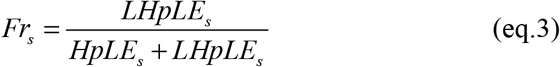

### 2.4. Statistical analysis: cancer and emotional happiness

First, we analyzed whether emotional happiness among the general panels differed by age and sex groups using a two-way analysis of variance of arcsine-transformed data. Subsequently, the validity of emotional happiness was confirmed by calculating the κ coefficients between emotional happiness and enjoyment or laughter.

We investigated whether cancer incidence decreased emotional happiness using data from the questionnaires for the general panel and the disease panel with cancer history. We excluded those who did not provide their health history (445 participants) in this analysis. First, we analyzed the association between cancer history and emotional happiness for each sex using a chi-square test. As in previous reports (Arai et al., 2020; Murakami et al., 2021; Murakami et al., 2018), we adjusted for covariates using propensity score matching (Rosenbaum and Rubin, 1983) to examine whether cancer incidence decreased emotional happiness for each sex. Covariates included subjective feelings of health (“not good at all, not good” or “good, fair, normal”), psychological distress (“K6 ≤ 12” or “K6 ≥13” (Kessler et al., 2003)), and health history, including hypertension, diabetes, hyperlipidemia (“yes” or “no,” respectively), age (“< 50 years,” “50–59,” or “≥ 60 years”), job (“company employee, civil servants and nonprofit organization employees, teachers, healthcare professionals, and other professionals,” “farmer, forestry, fishery workers, and other self-employed,” or “others”), marital status (“married, living together” “married, living separately” or “other”), presence or absence of each of children and grandchildren, educational background (“junior or high school graduate,” “university etc. graduate.” or “do not want to answer”), presence of a jobless person within the household (“presence,” “absence,” or “do not want to answer”), household income (“< 3 million yen,” “3–6 million yen” “≥ 6 million yen,” or “do not want to answer”), and smoking habit (“ smokes” or “does not smoke”). We used 1:1 nearest-neighbor matching with a ± 0.1 caliper and no replacement, as described previously (Murakami et al., 2018). We obtained 502 pairs of males (mean caliper: ± 0.0075) and 339 pairs of females (mean caliper: ± 0.00021). McNemar’s test was used to examine differences in emotional happiness between those with and without cancer for each sex after matching.

In addition, we used questionnaires administered to the disease panel with cancer history to examine the associations between emotional happiness and cancer type (lung, colon, stomach, prostate, breast, or other cancers), history (“< 2 years,” “2–4 years,” or “≥ 4 years”), or stage (“0,” “I,” “II,” “III or more,” or “do not know”), using the chi-square or Fisher’s exact test.

All analyses were performed using IBM SPSS Statistics 28 or R statistical software (R Development Core Team, 2021; Randolph et al., 2014).

### 2.5. Risk exposures

#### 2.5.1. Radon

Radon exposure in Japan was set at 15 Bq/m^3^ based on monitoring data from 1994 to 1996 (Sanada et al., 1999) and 2007 to 2010 (Suzuki et al., 2010) in a previous report (Nuclear Safety Research Association, 2020). We used a hazard ratio of 1.15 for lung cancer mortality per 100 Bq/m^3^ increase in radon concentration in the general United States population to establish the dose-response equation, assuming a linear relationship between dose and response (Turner et al., 2011). The relative risk was assumed to be independent of age and sex. Lung cancer mortality rates due to radon by age were calculated using previously reported age-specific lung cancer mortality rates for males and females in Japan (Statistics of Japan, 2021). We calculated LLE and LHpLE in the absence of radon by subtracting the mortality rate due to radon from the age-specific mortality rates in the life table because the Japanese life table (Ministry of Health Labour and Welfare, 2012) is based on all-cause mortality, including radon risk. We multiplied these values by − 1 to estimate the LLE and LHpLE due to radon exposure.

The analysis described in “section *2*.*4. Statistical analysis: cancer and emotional happiness*” demonstrated no significant decrease in emotional happiness due to cancer for males and females, and no significant associations between emotional happiness and cancer type, history, or stage among patients with cancer (see details in “*3*.*2. Association between cancer and emotional happiness*”). Emotional happiness was significantly higher in males with cancer than those without cancer; however, we considered it inappropriate to assume that cancer increased emotional happiness. Therefore, we did not consider the decrease in emotional happiness associated with cancer due to radon. The same applies to arsenic and PM_2.5_ in the following sections.

#### 2.5.2. Arsenic

The arsenic risk was calculated using the carcinogenic slope factor (CSF) or its drinking water equivalent. The endpoints were skin, liver, and lung cancers. The CSF for skin cancer was calculated using the median oral slope factor of 1.5 (mg/kg/day)^− 1^ as given by the United States Environmental Protection Agency’s Integrated Risk Information System (U.S. Environmental Protection Agency, 2002). The median CSF for liver and lung cancers were 0.00000306 (μg/L)^− 1^ and 0.0000385 (μg/L) ^− 1^, respectively (Health Canada, 2006). CSF was constant, regardless of age or sex.

The baseline cancer mortality rates used for life table analysis were obtained from a previous report (Statistics of Japan, 2021). Establishing a fatality rate was crucial for participants with skin cancer because the CSF for skin cancer (U.S. Environmental Protection Agency, 2002) represents incidence rates. Here, we used cancer incidence data from the Ministry of Health, Labour, and Welfare in 2010 (Cancer information service National Cancer Center Japan, 2022a; Hori et al., 2015). We also referred to the cancer incidence data from 2016 (Cancer information service National Cancer Center Japan, 2022b). These data were used to calculate the ratio of fatality to skin cancer cases: 0.08 in 2010 and 0.05 in 2016. Subsequently, a conservative estimate of 0.1 was used in this study.

We focused on inorganic arsenic and considered the sum of its oral intake from dietary sources and drinking water for arsenic exposure in Japan. We adopted 20 μg/day as the oral intake from dietary sources because the dietary intake of inorganic arsenic was 19.4 μg/day in a nationwide market basket survey in Japan (Akiyama et al., 2020). Drinking tap water was assumed to be the exposure route of oral intake of drinking water. The maximum arsenic concentration in tap water was 0.1 μg/L, with 91% of water purification plants having concentrations of 0.01 μg/L or less (Japan Water Works Association, 2021). Therefore, the maximum concentration scenario (Scenario A) was set at 0.1 μg/L, and the average concentration scenario (Scenario B) was set at 0.005 μg/L, which is half the quantification limit. The arsenic levels were assumed to be inorganic. The daily drinking water consumption was assumed to be 2 L/day. Assuming a body weight of 50 kg, the exposure levels were 0.0008 mg/kg/day (Scenario A) and 0.00042 mg/kg/day (Scenario B).

In the life table analysis, we converted CSFs to relative risks (World Health Organization Regional Office for Europe, 2000). Similar to the method used for radon, LLE and LHpLE without arsenic were calculated and multiplied by − 1 to obtain LLE and LHpLE due to arsenic since the Japanese life table (Ministry of Health Labour and Welfare, 2012) includes arsenic risk. LLE and LHpLE were calculated for skin cancer only and all skin, liver, and lung cancers.

#### 2.5.3. PM_2.5_

An epidemiological study on PM_2.5_ in Japan reported a significant association between PM_2.5_ exposure and increased mortality from lung cancer (Katanoda et al., 2011). Therefore, we used lung cancer as the risk indicator endpoint. We also used data from another large epidemiological study comprising 1.2 million people with 16 years of follow-up in the United States (Pope III et al., 2002). The LHpLE calculations using each relative risk are hereafter referred to as scenarios X and Y. The cutoff values were 15 and 10 μg/m^3^ for scenarios X and Y, respectively. The risk increased by a factor of 1.24 and 1.08, respectively, for each 10 μg/m^3^ increase in exposure concentration above the cutoff value. The relative risk was assumed to be independent of age and sex. The baseline mortality rates of lung cancer used for life table analysis were obtained from a previous report (Statistics of Japan, 2021).

Exposure concentrations were obtained from the national data for 2012 and 2020 from 596 and 1119 measurement sites, respectively (National Institute for Environmental Studies, 2022). During this period, a large decrease in PM_2.5_ concentration was observed in Japan (from 15.2 to 9.59 μg/m^3^). In this study, 2012 was chosen because of the small number of measurement sites before 2011, which was insufficient for calculating representative values of exposure concentrations in Japan. The risk reductions due to the improvements in air quality were evaluated for these two time points. A histogram of exposure concentrations at 1 μg/m^3^ intervals was created for each prefecture. The population distribution of exposure concentrations by prefecture was obtained by weighting the population by the frequency of the histograms. Population data by prefecture were obtained from national data (e-Stat, 2022). This was performed for all 47 prefectures and summed to obtain the population distribution of exposure concentrations in Japan. The relative risk corresponding to the exposure concentration was determined. The relative risk for each exposure concentration was multiplied by the corresponding population and weighted and averaged to obtain a representative value of the relative risk. In scenario X, the relative risks were 1.0250 and 1.0002 for exposure concentrations in 2012 and 2020, respectively. The relative risks were 1.0377 and 1.0030, respectively, for scenario Y. These relative risks were used for life-table analysis.

LLE and LHpLE without PM_2.5_ were calculated and multiplied by − 1 to obtain LLE and LHpLE due to PM_2.5_ because the Japanese life table (Ministry of Health Labour and Welfare, 2012) includes life expectancy due to PM_2.5_ risk.

#### 2.5.4. Psychological distress

LHpLE due to psychological distress was analyzed using K6. In line with a previous report (Kessler et al., 2003), a K6 score of 13 points or higher was considered a risk factor associated with serious mental illness. The K6 values for the Japanese population were based on a national survey in 2019 (Ministry of Health Labour and Welfare, 2020). The percentage with a K6 of 13 points or higher (*P*_*x,s*_) was calculated for each age and sex group. We assumed a uniform distribution over the range of 10–14 points: two-fifths of the people scoring 10–14 would have scored 13–14 since data were presented on a 5-point scale, as in a previous report (Murakami et al., 2018). We assumed that the percentage with a K6 score of 13 points or higher for participants aged 11 and younger was the same as that for individuals aged 12–19. This study did not consider the increase in mortality due to psychological distress, although it can increase mortality (Pratt, 2009). We used ∆*Hp*_*s*_ (− 0.19 for males and − 0.22 for females), representing the reduced emotional happiness due to psychological distress (Murakami et al., 2018). LHpLE in the absence of psychological distress was determined by calculating the emotional happiness *Hp*_*x,s*_ among the population in the absence of psychological distress using eq. 4, and multiplying this value by − 1 to obtain LHpLE due to psychological distress.

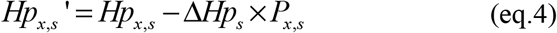

## 3. Results

### 3.1. Emotional happiness by age and sex groups

Emotional happiness by age and sex groups varied, ranging from 0.43 to 0.50 for males in their 20s to 60s and 0.51 to 0.62 for females in their 20s to 60s (Figure 1). The interactions between age and sex groups were insignificant (*P* > 0.05), but significant differences were observed between age and sex groups (*P* < 0.001 for each). A significant positive association was observed between emotional happiness and enjoyment or laughter, with κ coefficients of 0.81 and 0.42, respectively (*P* < 0.001). HpLE calculated using this emotional happiness was 37.5 years and 49.1 years for males and females, respectively.

**Figure 1.**
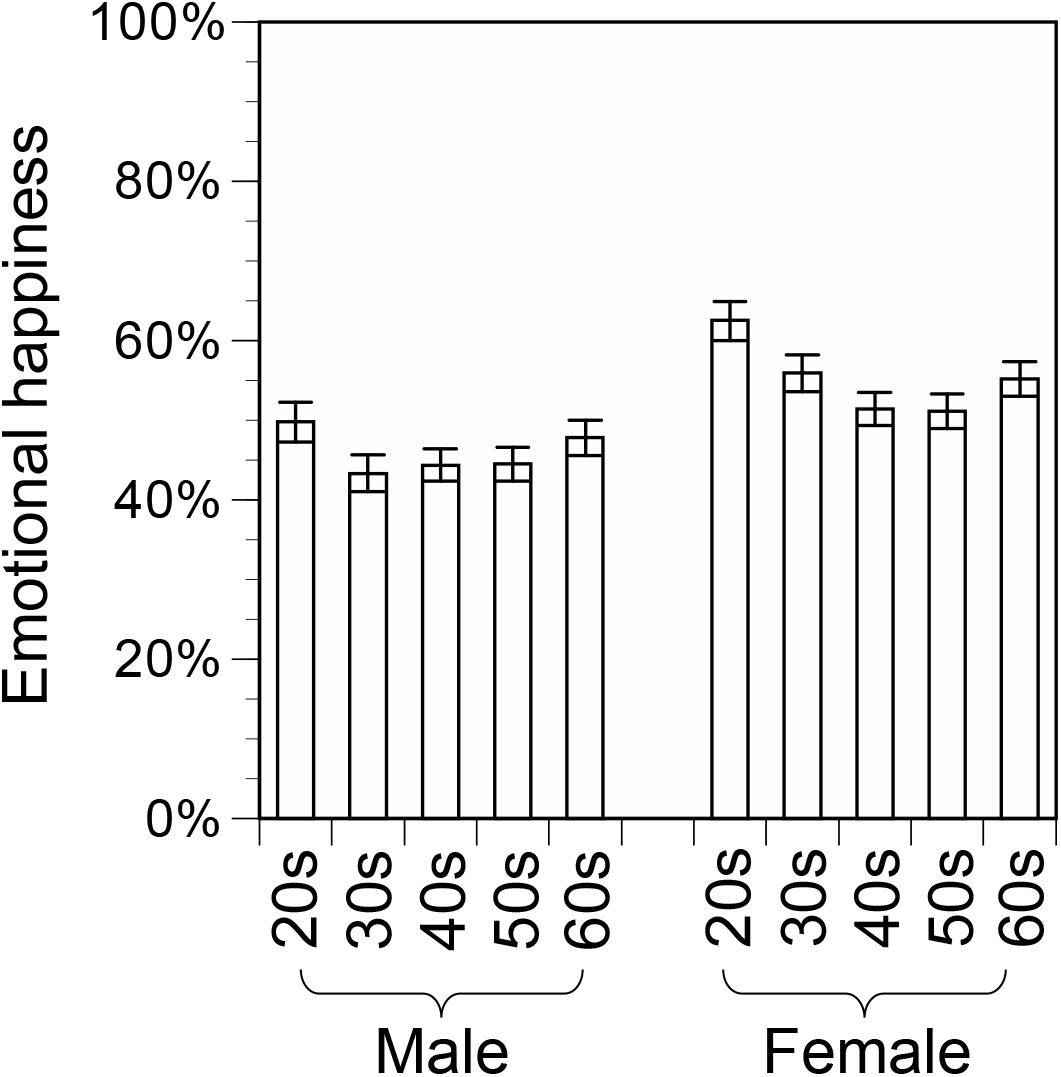
Emotional happiness in the general panel by age and sex groups. The error bar represents a standard error. Two-way analysis of variance of arcsine-transformed data demonstrated no significant interaction between age and sex (*P* > 0.05), but significant differences were observed between age groups (*P* < 0.001) and sex (*P* < 0.001).

### 3.2. Association between cancer and emotional happiness

The chi-square test revealed no significant difference in emotional happiness between females with and without cancer (*P* > 0.05); however, among males, patients with cancer had significantly higher emotional happiness than patients without cancer (*P* < 0.001) (Figure 2). No significant difference in emotional happiness was observed between females with and without cancer (*P* > 0.05), whereas males with cancer had significantly higher emotional happiness than those without cancer (*P* = 0.01, ∆Hp = 0.078) after adjusting for covariates using propensity score matching.

**Figure 2.**
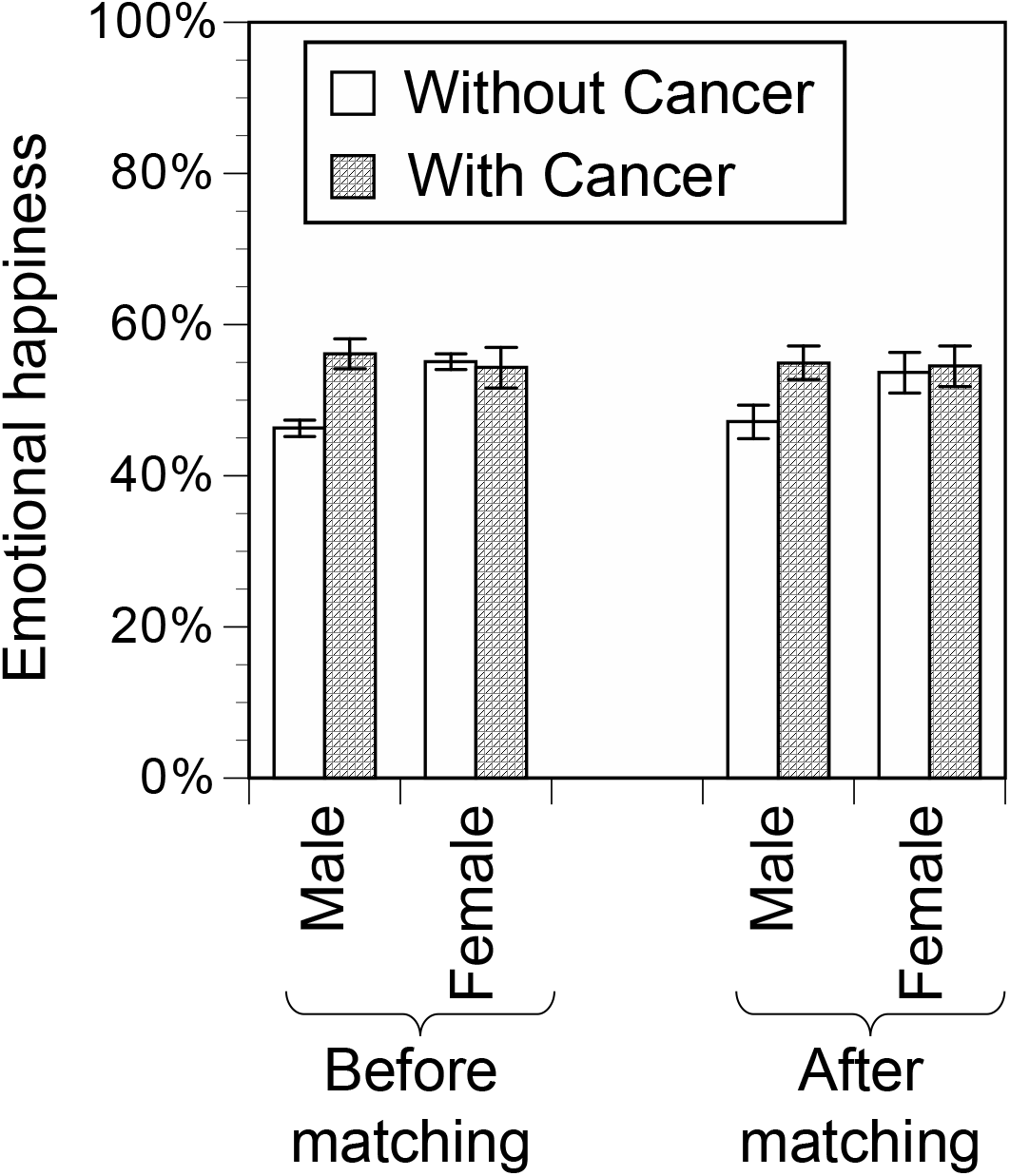
Comparisons of emotional happiness between the absence and presence of cancer before and after propensity score matching. The error bar represents a standard error. The chi-square test before matching and the McNemar test after matching demonstrated insignificant differences in emotional happiness between the absence and presence of cancer in females (*P* > 0.05). These tests showed that emotional happiness in males with cancer was significantly higher than in those without cancer (*P* < 0.001 before matching; *P* = 0.01 after matching). For matching, covariates were adjusted for subjective feelings of health, health history (hypertension, diabetes, and hyperlipidemia), psychological distress, age, job, marriage, child, grandchild, educational background, presence of a jobless person within the household, household income, and smoking habit.

No significant associations were observed between emotional happiness and cancer type (lung, colorectal, stomach, prostate, breast, and other cancers), history, or stage (*P* > 0.05) (Table 1).

**Table 1.** Associations between emotional happiness and attributes of patients with cancer.

|  | Male |  |  | Female |  |  |
| --- | --- | --- | --- | --- | --- | --- |
|  | Emotional<br>happiness (+) | Emotional<br>happiness (–) | *P* | Emotional<br>happiness (+) | Emotional<br>happiness (–) | *P* |
| Lung cancer (+) | 41 (55%) | 34 (45%) | 0.61 | 5 (36%) | 9 (64%) | 0.14 |
| Lung cancer (–) | 296 (58%) | 216 (42%) |  | 138 (56%) | 108 (44%) |  |
| Colorectal cancer (+) | 78 (57%) | 60 (43%) | 0.81 | 15 (52%) | 14 (48%) | 0.71 |
| Colorectal cancer (–) | 259 (58%) | 190 (42%) |  | 128 (55%) | 103 (45%) |  |
| Stomach cancer (+) | 57 (56%) | 45 (44%) | 0.73 | 4 (57%) | 3 (43%) | 1.00 |
| Stomach cancer (–) | 280 (58%) | 205 (42%) |  | 139 (55%) | 114 (45%) |  |
| Prostate cancer (+) | 76 (57%) | 58 (43%) | 0.85 | 0 (–) | 0 (–) | – |
| Prostate cancer (–) | 261 (58%) | 192 (42%) |  | 143 (55%) | 117 (45%) |  |
| Breast cancer (+) | 3 (38%) | 5 (63%) | 0.30 | 70 (55%) | 58 (45%) | 0.92 |
| Breast cancer (–) | 334 (58%) | 245 (42%) |  | 73 (55%) | 59 (45%) |  |
| Other cancer (+) | 157 (58%) | 115 (42%) | 0.89 | 59 (55%) | 48 (45%) | 0.97 |
| Other cancer (–) | 180 (57%) | 135 (43%) |  | 84 (55%) | 69 (45%) |  |
| Cancer history: < 2 years | 42 (49%) | 44 (51%) | 0.09 | 17 (50%) | 17 (50%) | 0.48 |
| Cancer history: 2–4 years | 142 (62%) | 87 (38%) |  | 50 (60%) | 33 (40%) |  |
| Cancer history: ≥ 4 years | 153 (56%) | 120 (44%) |  | 76 (53%) | 67 (47%) |  |
| Stage 0 | 53 (62%) | 32 (38%) | 0.85 | 30 (68%) | 14 (32%) | 0.13 |
| Stage I | 93 (56%) | 73 (44%) |  | 49 (58%) | 35 (42%) |  |
| Stage II | 67 (57%) | 51 (43%) |  | 28 (53%) | 25 (47%) |  |
| Stage III or more | 70 (56%) | 55 (44%) |  | 22 (44%) | 28 (56%) |  |
| Do not know | 53 (60%) | 35 (40%) |  | 12 (44%) | 15 (56%) |  |

### 3.3. LLE and LHpLE due to environmental carcinogenic chemicals and psychological distress

Table 2 compares the LLE due to environmental carcinogenic chemicals with those in previous studies conducted in Japan (Gamo et al., 2003; GBD 2019 Risk Factors Collaborators, 2020; Institute for Health Metrics and Evaluation, 2015). The LLE due to radon calculated in this study differed from the previous report (Gamo et al., 2003) within about a factor of two, despite differences in exposure data and dose-response equations. The calculated LLE for arsenic in scenario B was 6.9 × 10^−4^ years for skin cancer only and 5.1 × 10^−3^ years for all endpoints (skin, liver, and lung cancers). LLE from arsenic-related skin cancer in a previous report (Gamo et al., 2003) was 1.6 × 10^−3^ years, and the difference between this study and the previous study was about a factor of two. LLEs calculated in this study varied greatly in years for PM_2.5_, depending on the dose-response equation and the baseline values: for exposure in 2012, LLE was 1.4 × 10^−2^ years in the epidemiological study for Japan (Scenario X) (Katanoda et al., 2011) and 2.2 × 10^−2^ years in the epidemiological study in the United States (Scenario Y) (Pope III et al., 2002) and 2020 exposure were 9.7 × 10^−5^ years and 1.7 × 10^−3^ years, respectively. In Japan, LLE from lung cancer caused by PM_2.5_ calculated by the Global Burden Disease 2019 was 9.6 × 10^−4^ years (GBD 2019 Risk Factors Collaborators, 2020; Institute for Health Metrics and Evaluation, 2015). The LLEs for scenarios X and Y were approximately one-tenth and two times higher, respectively, than those previously reported.

**Table 2.**
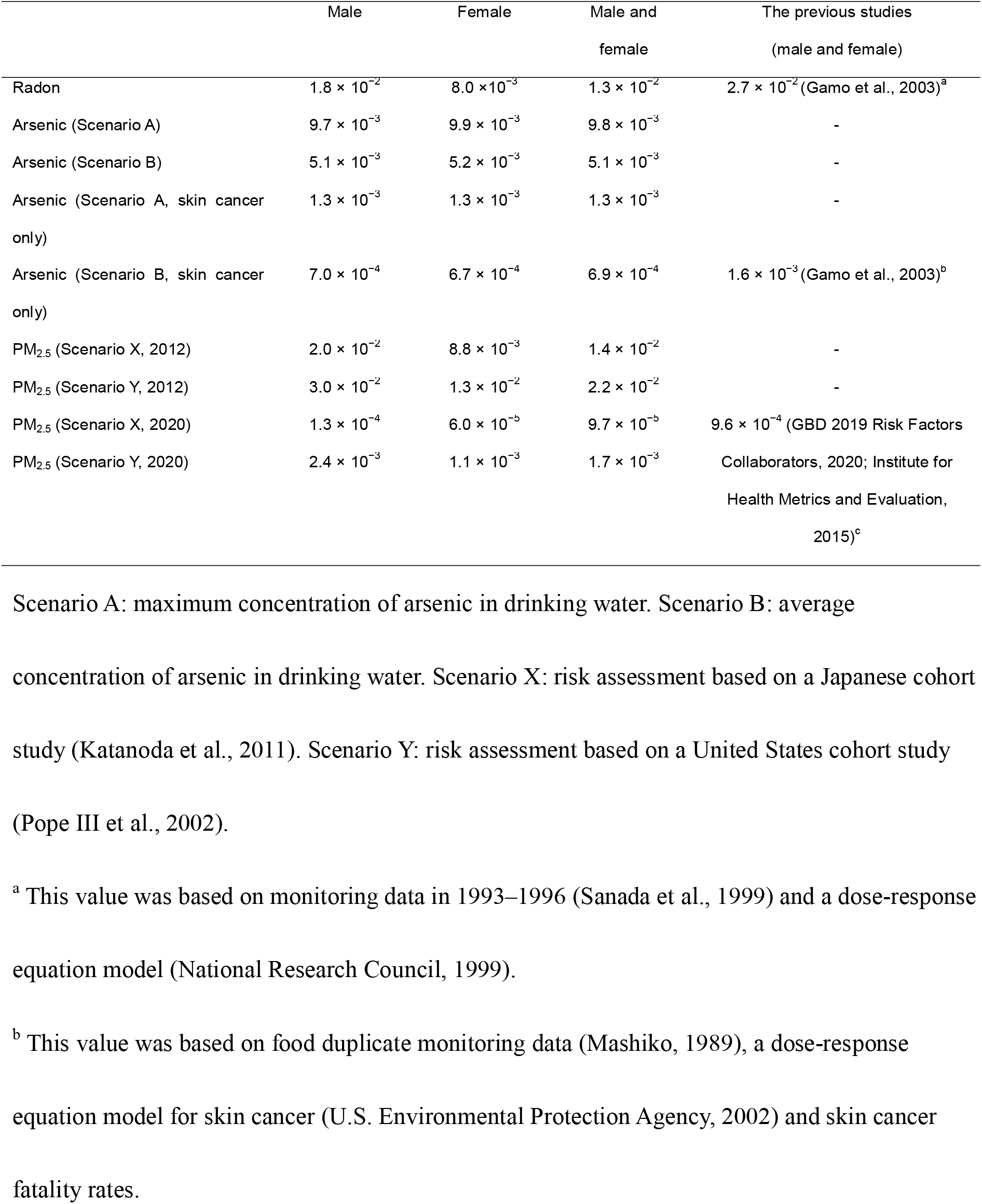
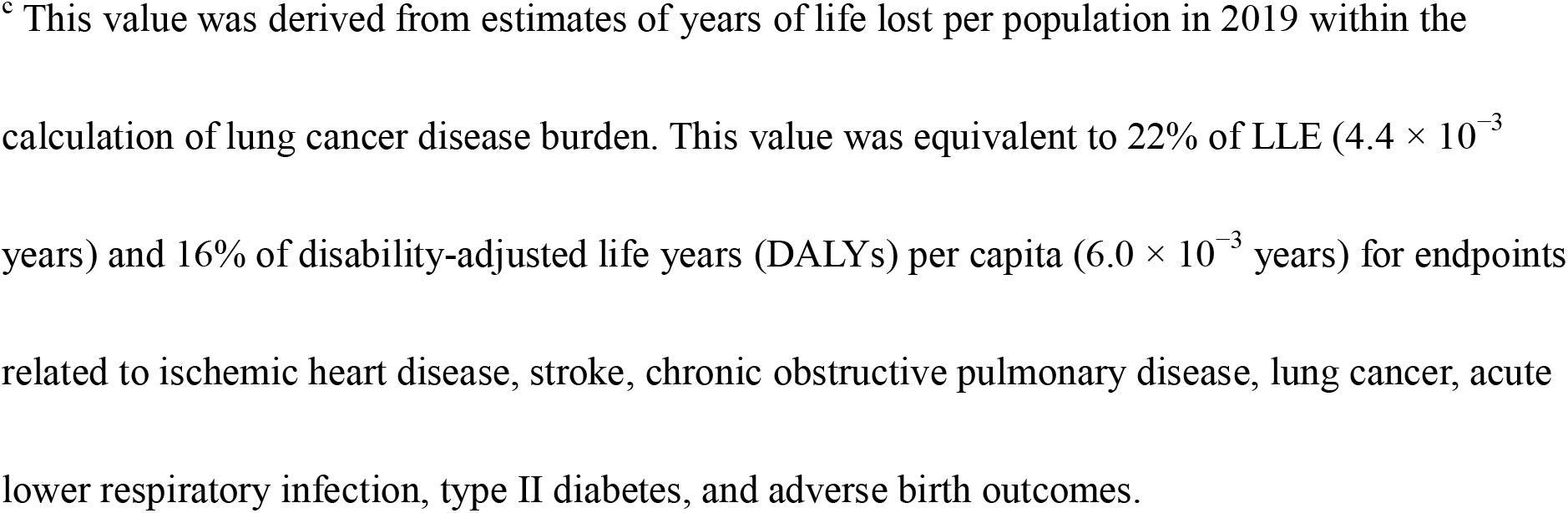
Loss of life expectancy (LLE) due to radon, arsenic, and PM_2.5_ in Japan and comparison with previous studies [years].

|  | Male | Female | Male and female | The previous studies (male and female) |
| --- | --- | --- | --- | --- |
| Radon | $1.8 \times 10^{-2}$ | $8.0 \times 10^{-3}$ | $1.3 \times 10^{-2}$ | $2.7 \times 10^{-2}$ (Gamo et al., 2003)<sup>a</sup> |
| Arsenic (Scenario A) | $9.7 \times 10^{-3}$ | $9.9 \times 10^{-3}$ | $9.8 \times 10^{-3}$ | - |
| Arsenic (Scenario B) | $5.1 \times 10^{-3}$ | $5.2 \times 10^{-3}$ | $5.1 \times 10^{-3}$ | - |
| Arsenic (Scenario A, skin cancer only) | $1.3 \times 10^{-3}$ | $1.3 \times 10^{-3}$ | $1.3 \times 10^{-3}$ | - |
| Arsenic (Scenario B, skin cancer only) | $7.0 \times 10^{-4}$ | $6.7 \times 10^{-4}$ | $6.9 \times 10^{-4}$ | $1.6 \times 10^{-3}$ (Gamo et al., 2003)<sup>b</sup> |
| PM<sub>2.5</sub> (Scenario X, 2012) | $2.0 \times 10^{-2}$ | $8.8 \times 10^{-3}$ | $1.4 \times 10^{-2}$ | - |
| PM<sub>2.5</sub> (Scenario Y, 2012) | $3.0 \times 10^{-2}$ | $1.3 \times 10^{-2}$ | $2.2 \times 10^{-2}$ | - |
| PM<sub>2.5</sub> (Scenario X, 2020) | $1.3 \times 10^{-4}$ | $6.0 \times 10^{-5}$ | $9.7 \times 10^{-5}$ | $9.6 \times 10^{-4}$ (GBD 2019 Risk Factors Collaborators, 2020; Institute for Health Metrics and Evaluation, 2015)<sup>c</sup> |
| PM<sub>2.5</sub> (Scenario Y, 2020) | $2.4 \times 10^{-3}$ | $1.1 \times 10^{-3}$ | $1.7 \times 10^{-3}$ | |

Scenario A: maximum concentration of arsenic in drinking water. Scenario B: average concentration of arsenic in drinking water. Scenario X: risk assessment based on a Japanese cohort study (Katanoda et al., 2011). Scenario Y: risk assessment based on a United States cohort study (Pope III et al., 2002).
<sup>a</sup> This value was based on monitoring data in 1993–1996 (Sanada et al., 1999) and a dose-response equation model (National Research Council, 1999).
<sup>b</sup> This value was based on food duplicate monitoring data (Mashiko, 1989), a dose-response equation model for skin cancer (U.S. Environmental Protection Agency, 2002) and skin cancer fatality rates.
<sup>c</sup> This value was derived from estimates of years of life lost per population in 2019 within the calculation of lung cancer disease burden. This value was equivalent to 22% of LLE ( $4.4 \times 10^{-3}$ years) and 16% of disability-adjusted life years (DALYs) per capita ( $6.0 \times 10^{-3}$ years) for endpoints related to ischemic heart disease, stroke, chronic obstructive pulmonary disease, lung cancer, acute lower respiratory infection, type II diabetes, and adverse birth outcomes.

Table 3 compares LHpLE due to environmental carcinogenic chemicals with that due to psychological distress. The average LHpLE for males and females was 6.4 × 10^−3^ years for radon, 5.0 × 10^−3^ years (Scenario A, all endpoints) and 2.6 × 10^−3^ years (Scenario B, all endpoints) for arsenic, 7.2 × 10^−3^ years (Scenario X) and 1.1 × 10^−2^ years (Scenario Y) for PM_2.5_ exposure in 2012, and 4.8 × 10^−5^ years (Scenario X) and 8.6 × 10^−4^ years (Scenario Y) for PM_2.5_ exposure in 2020. In contrast, LHpLE due to psychological distress was 9.7 × 10^−1^ years. *Fr* was 1.5 × 10^−4^, 1.2 × 10^−4^, 6.1 × 10^−5^, 1.7 × 10^−4^, 2.5 × 10^−4^, 1.1 × 10^−6^, 2.0 × 10^−5^, and 2.2 × 10^−2^, respectively.

**Table 3.** Comparison of the loss of happy life expectancy (LHpLE) among radon, arsenic, PM_2.5_, and psychological distress in Japan [years].

|  | Male | Female | Male and female |
| --- | --- | --- | --- |
| Radon | $8.5 \times 10^{-3}$ ($2.3 \times 10^{-4}$) | $4.4 \times 10^{-3}$ ($8.9 \times 10^{-5}$) | $6.4 \times 10^{-3}$ ($1.5 \times 10^{-4}$) |
| Arsenic (Scenario A) | $4.6 \times 10^{-3}$ ($1.2 \times 10^{-4}$) | $5.4 \times 10^{-3}$ ($1.1 \times 10^{-4}$) | $5.0 \times 10^{-3}$ ($1.2 \times 10^{-4}$) |
| Arsenic (Scenario B) | $2.4 \times 10^{-3}$ ($6.5 \times 10^{-5}$) | $2.8 \times 10^{-3}$ ($5.8 \times 10^{-5}$) | $2.6 \times 10^{-3}$ ($6.1 \times 10^{-5}$) |
| Arsenic (Scenario A, skin cancer only) | $6.3 \times 10^{-4}$ ($1.7 \times 10^{-5}$) | $7.0 \times 10^{-4}$ ($1.4 \times 10^{-5}$) | $6.7 \times 10^{-4}$ ($1.5 \times 10^{-5}$) |
| Arsenic (Scenario B, skin cancer only) | $3.3 \times 10^{-4}$ ($8.9 \times 10^{-6}$) | $3.7 \times 10^{-4}$ ($7.5 \times 10^{-6}$) | $3.5 \times 10^{-4}$ ($8.1 \times 10^{-6}$) |
| PM<sub>2.5</sub> (Scenario X, 2012) | $9.5 \times 10^{-3}$ ($2.5 \times 10^{-4}$) | $4.9 \times 10^{-3}$ ($9.9 \times 10^{-5}$) | $7.2 \times 10^{-3}$ ($1.7 \times 10^{-4}$) |
| PM<sub>2.5</sub> (Scenario Y, 2012) | $1.4 \times 10^{-2}$ ($3.8 \times 10^{-4}$) | $7.3 \times 10^{-3}$ ($1.5 \times 10^{-4}$) | $1.1 \times 10^{-2}$ ($2.5 \times 10^{-4}$) |
| PM<sub>2.5</sub> (Scenario X, 2020) | $6.4 \times 10^{-5}$ ($1.7 \times 10^{-6}$) | $3.3 \times 10^{-5}$ ($6.7 \times 10^{-7}$) | $4.8 \times 10^{-5}$ ($1.1 \times 10^{-6}$) |
| PM<sub>2.5</sub> (Scenario Y, 2020) | $1.1 \times 10^{-3}$ ($3.0 \times 10^{-5}$) | $5.8 \times 10^{-4}$ ($1.2 \times 10^{-5}$) | $8.6 \times 10^{-4}$ ($2.0 \times 10^{-5}$) |
| Psychological distress | $7.5 \times 10^{-1}$ ($2.0 \times 10^{-2}$) | $1.2 \times 10^0$ ($2.4 \times 10^{-2}$) | $9.7 \times 10^{-1}$ ($2.2 \times 10^{-2}$) |

The values in parentheses represent the fraction of losses caused by risk exposure to HpLE.
Scenario A: maximum concentration of arsenic in drinking water. Scenario B: average concentration of arsenic in drinking water. Scenario X: risk assessment based on a Japanese cohort study (Katanoda et al., 2011). Scenario Y: risk assessment based on a United States cohort study (Pope III et al., 2002).

## 4. Discussion

In this study, we conducted questionnaire surveys among a general panel and disease panel with cancer history to investigate emotional happiness based on age and sex and analyzed the relationship between cancer incidence and emotional happiness. In addition, we compared LHpLE among carcinogenic chemicals, such as radon, arsenic, and PM_2.5_, and psychological distress in Japan.

Emotional happiness was significantly higher in females than in males and was positively associated with enjoyment and laughter, which are positive wellbeing items similar to emotional wellbeing. These results are consistent with those of a previous study (Murakami et al., 2018). Emotional happiness scores ranged from 0.43–0.50 for males in their 20s to 60s and 0.51–0.62 for females in their 20s to 60s, which was slightly lower than those reported in the 2015 study (0.47– 0.58 for males and 0.65–0.68 for females) (Murakami et al., 2018) and the 2016 study (0.52–0.62 for males) (Arai et al., 2020) conducted in Japan. Emotional happiness might reduce during the coronavirus disease 2019 outbreak, and the HpLE obtained in this study was possibly lower than it would be in the absence of the outbreak.

In this study, regardless of propensity score matching, emotional happiness was significantly higher in males with cancer than those without cancer. However, emotional happiness between females with cancer and those without cancer did not differ significantly. One possible reason for the increased emotional happiness among males with cancer is that their values might have changed following their cancer diagnosis. According to socioemotional selectivity theory, the perception that “time is finite” alters life goals, cognitions, and emotions, leading to a greater emphasis on emotional satisfaction (Carstensen, 2006). Cancer might enhance emotional happiness by reinforcing the perception of time as finite. However, we considered it inappropriate to conclude that cancer incidence increased emotional happiness when LHpLE was calculated. We did not consider the decrease or increase in emotional happiness due to cancer incidence when calculating LHpLE because no significant associations were observed between emotional happiness and cancer type, history, or stage.

In this study, as a preliminary step in discussing the magnitude of LHpLE for carcinogenic chemicals, we compared the LLE values with those reported in previous studies (Gamo et al., 2003; GBD 2019 Risk Factors Collaborators, 2020; Institute for Health Metrics and Evaluation, 2015). Regardless of the differences in exposure doses, dose-response equations, and life tables used, the differences from the previous study (Gamo et al., 2003) for radon and arsenic (skin cancer) were within a factor of two. However, when liver and lung cancers, which were not included in the previous study (Gamo et al., 2003), were considered for arsenic, the LLE was 7.5 times greater than that for skin cancer alone. Regarding PM_2.5_, owing to differences in baseline and dose-response equations from the different epidemiological studies used (Katanoda et al., 2011; Pope III et al., 2002), the LLE at 2020 exposure levels was 9.7 × 10^−5^ years (Scenario X) and 1.7 × 10^−3^ years (Scenario Y) in this study, yielding an 18-fold difference between the scenarios. In contrast, the LLE from lung cancer due to PM_2.5_ exposure in a previous study (GBD 2019 Risk Factors Collaborators, 2020; Institute for Health Metrics and Evaluation, 2015) was 9.6 × 10^−4^ years. These findings suggest that appropriate dose-response equations and baseline settings are important in risk assessments, especially in Japan, where atmospheric PM_2.5_ concentrations have been declining (National Institute for Environmental Studies, 2022). In general, it is desirable to use epidemiological data from the target region. However, scenario X is based on an epidemiological study conducted in Japan (Katanoda et al., 2011) with 63,520 participants and a study period of 10 years, which were smaller than those in the epidemiological study in the United States for scenario Y (1.2 million participants, 16 years) (Pope III et al., 2002). This may have resulted in a higher calculated baseline value owing to weak statistical power. Comparing the value from the 2019 Global Burden Disease, which had a dose-response equation and baseline value set for various countries (GBD 2019 Risk Factors Collaborators, 2020; Institute for Health Metrics and Evaluation, 2015), scenario X possibly underestimated the risk of PM_2.5_. Therefore, we refer to the values of scenario Y concerning the LHpLE for PM_2.5_.

The average LHpLE for males and females were 6.4 × 10^−3^ years for radon and 2.6 × 10^−3^ years for arsenic (Scenario B, all endpoints), 1.1 × 10^−2^ for PM_2.5_ at 2012 exposure levels, and 8.6 × 10^−4^ for PM_2.5_ at 2020 exposure levels (Scenario Y). LHpLE for PM_2.5_ decreased significantly from 2012 to 2020. Fr was 1.5 × 10^−4^, 6.1 × 10^−5^, 2.5 × 10^−4^, and 2.0 × 10^−5^, respectively. LHpLE due to radon, arsenic (Scenario B), and PM_2.5_ in 2012 (Scenario Y) were comparable to or larger than those due to radiation exposure after the Fukushima Daiichi Nuclear Power Station accident (5 × 10^−3^ for 40-year-old males and females) (Murakami et al., 2018). Given that the Fr value exceeded 10^−5^, environmental policies must reduce the risk of these carcinogenic chemicals, including PM_2.5_, whose concentration in the air has improved in recent years (National Institute for Environmental Studies, 2022). In this study, PM_2.5_ calculation only considered the risk associated with lung cancer; the LLE from lung cancer due to PM_2.5_ by the 2019 Global Burden Disease was equivalent to 22% of LLE (4.4 × 10^−3^ years) and 16% of DALYs per capita (6.0 × 10^−3^ years) for all endpoints related to ischemic heart disease, stroke, chronic obstructive pulmonary disease, lung cancer, acute lower respiratory infection, type II diabetes, and adverse birth outcomes (GBD 2019 Risk Factors Collaborators, 2020; Institute for Health Metrics and Evaluation, 2015). The LHpLE due to PM_2.5_ could be nearly an order of magnitude higher than that calculated in this study if various endpoints other than lung cancer were included since lung cancer partly contributes to all endpoints in the 2019 Global Burden Disease.

The LHpLE associated with environmental carcinogenic chemicals were approximately two to four orders of magnitude lower than that due to psychological distress. However, this does not mean that policies targeting psychological distress should be prioritized over carcinogen risk management. For example, assessing the cost-effectiveness of the measures for each risk exposure is essential for policymaking. In addition, the LHpLE used in this study can evaluate the effectiveness of mitigation measures for different risks in various fields in the future. The LHpLE indicator can be beneficial in how risk-reduction decisions are made and how the effects are achieved by comparing the magnitude and cost-effectiveness of these various risks.

This study had several limitations. First, this study used online surveys to calculate the values of emotional happiness for the general population and patients with cancer. The target population may differ from that of Japan as a whole. However, our online surveys had the advantage of reducing bias because participants were incentivized to participate by being awarded points, even if they were uninterested in the survey content. In addition, the participants in this study were recruited to match the age and sex distributions of the whole country to reduce the impact of selection bias on emotional happiness. However, this was applied only to the general panel and not to the cancer panel, which had a limited sample size. Furthermore, to discuss changes in emotional happiness due to cancer, the participants were matched for various covariates, such as educational background, annual income, and health history, to reduce the impact of selection bias. Second, we substituted values for emotional happiness for those under 20 and over 70 years of age with values for those in their 20s and 60s, respectively. However, as shown in the sensitivity analysis of a previous study (Murakami et al., 2018), the impact of this assumption on LHpLE can be negligibly small. Third, this study did not consider the increase in mortality due to psychological distress. Similarly, we did not consider the risk for those with moderate psychological distress between 5 and 12 points on the K6 scale. Thus, the risk of psychological distress may have been underestimated. Fourth, as noted above, there were large differences in the risk of PM_2.5_ depending on the dose-response equation, baseline value, and endpoints applied. Updated findings based on sophisticated, large-scale cohort studies in the target regions are required to reduce the uncertainty in the PM_2.5_ risk assessment. Nevertheless, this study emphasized the importance of reducing PM_2.5_ risk.

## Data Availability

We have included all the data produced in the present work in the manuscript. The data used in the present study are available upon reasonable request to the authors.

## 5. Conclusions

This study compared the risk of radon, arsenic, and PM_2.5_ with that of psychological distress in Japan using the LHpLE indicator. The findings of this study are as follows.

- No significant reduction was observed in emotional happiness owing to cancer incidence, and no associations were observed between emotional happiness and cancer type, history, or stage.
- LHpLE was 6.4 × 10^−3^ years for radon, 2.6 × 10^−3^ years for arsenic (scenario B), 1.1 × 10^−2^ years for PM_2.5_ at 2012 exposure levels (Scenario Y), and 8.6 × 10^−4^ years at 2020 exposure level (Scenario Y). Fr exceeded 10^−5^, suggesting that risk reduction for these chemicals is important in environmental policies.
- LLE and LHpLE for PM_2.5_ at 2020 exposure levels differed greatly depending on the dose-response equation and baseline values. This warrants further research to obtain updated knowledge based on sophisticated, large-scale cohort studies in the target regions under low atmospheric PM_2.5_ concentrations, such as in Japan, to reduce the uncertainty of PM_2.5_ risk assessment.
- LHpLE due to radon, arsenic, and PM_2.5_ were two to four orders of magnitude lower than that due to psychological distress.
- LHpLE allowed the comparison of risks in various fields, including environmental chemicals and psychological distress, highlighting the strength of using the LHpLE indicator to compare the magnitude of risk and cost-effectiveness for various future risk exposures, thereby contributing to effective risk reduction decision-making.

## Acknowledgements

This work was supported by JSPS KAKENHI (grant number JP20H04354) and “The Nippon Foundation - Osaka University Project for Infectious Disease Prevention.”

